# Factors Associated with Intention to Vaccinate against COVID-19 in Puerto Rico

**DOI:** 10.1101/2021.03.19.21253972

**Authors:** Kyle Melin, Cheyu Zhang, Juan Pablo Zapata, Yonaira M. Rivera, Katie Fernandez, Enbal Shacham, Souhail M. Malavé-Rivera, Carlos E. Rodríguez-Díaz

## Abstract

COVID-19 has been particularly devastating to Black and Latinx communities in the U.S. However, data on acceptability of the COVID-19 vaccines among minority populations are limited. We conducted an online survey among adults in Puerto Rico to identify factors associated with intention to vaccinate against COVID-19. Sociodemographic variables were analyzed independently for association with intention to vaccinate. Significant associations were included in the multivariate logistic regression analysis. A total of 1016 responses were available for analysis. In the bivariate analysis, younger age, higher education, pre-covid employment, male sex, gay/bisexual identity, and single marital status were associated with increased intention to vaccinate. In the multivariate logistic regression, younger, male respondents who had higher educational attainment reported higher intention to vaccinate. Lower-income and living outside the San Juan metro region were associated with lower intention to vaccinate. National and international health organizations were identified as the most reliable sources of information, followed by healthcare professionals. These findings highlight the importance of considering sociodemographic characteristics identified with low intention to vaccinate as well as using trusted sources of information when designing public messaging related to increasing COVID-19 vaccinations.

## Introduction

As of this writing, the United States Centers for Disease Control and Prevention (CDC) has recommended the use of two COVID-19 vaccines, with more in development. Vaccination campaigns for COVID-19 will be key in curbing this global pandemic but must be purposeful to ensure they do not propagate existing disparities of COVID-19’s impact on communities of color.

COVID-19 has been particularly devastating to Black and Latino/Hispanic (henceforth: Latinx) communities in the U.S.^1,2^ Long-standing social inequities in social determinants of health place communities of color at elevated risk of COVID-19 infection, severe illness, and death compared to non-Hispanic whites.^3^ Therefore, it is imperative to document and strategize how COVID-19 vaccines can be successfully disseminated throughout these communities.

Previous research shows that Latinx adults are less likely than non-Hispanic whites to be vaccinated.^4,5^ Low perceived risk of influenza and healthcare providers’ failure to recommend vaccination may contribute to lower vaccination rates.^6^ Compared to non-Hispanic whites, U.S. Latinxs reported more vaccine-related side effects and were more likely to be misinformed about the availability of vaccines.^7^ In addition to individual-level barriers, systemic barriers have been well documented. Latinxs in the U.S. are less likely to have access to health care and more likely to suffer the effects of discriminatory policies, higher healthcare costs, lack of insurance, and language barriers; all of these factors constitute significant challenges to equitable public health efforts.^2^

Data on the acceptability of the COVID-19 vaccines among minority populations are limited. However, a cross-sectional survey of a representative sample of U.S. adults found that both Black and Latinx participants reported lower intent to be vaccinated for COVID-19 compared to non-Hispanic whites.^8^ Another nationally representative longitudinal study found that intention to vaccinate declined between April and December of 2020, with the highest decline observed among U.S. Hispanics.^9^ This is consistent with previous findings from the 2009 H1N1 pandemic which observed that intention to vaccinate may be highest early in a pandemic before a vaccine is available.^10^

In this research brief, we report the factors associated with the intention to vaccinate against COVID-19 among adults living in Puerto Rico.

## Methods

We conducted an online survey among adults (>21yo) living in Puerto Rico to identify social and behavioral factors impacted by the COVID-19 pandemic. The data collection instrument was available in Spanish (Puerto Rico’s official language), and several measures to assess socio-demographics, participants’ experiences with COVID-19 information sources, perceived impact of COVID-19, and perceived governmental response were culturally adapted following a process previously used by the research team.^11,12^ Intention to get vaccinated against COVID-19 was assessed asking “If there existed an approved vaccine to prevent the coronavirus infection, how likely is it that you would get vaccinated?” followed by a 4-point scale (not at all likely to very likely). All measured variables were analyzed independently for association with intention to vaccinate. Variables with significant association were included in the multivariate logistic regression analysis.

Participants were recruited using social media platforms and with the support of several community-based organizations that sent blast emails and posted study information on their websites and social media from July 7 to July 13, 2020. After screening and providing consent to participate, a total of n=1031 participants completed the survey. All research procedures were approved by the Human Research Subjects Protection Office of HIDDEN FOR PEER REVIEW.

## Results

A total of 1016 survey responses were available for analysis from respondents who reported their intention to get vaccinated for COVID-19. Respondents had a broad distribution of demographic and socioeconomic characteristics (Table 1), with a mean age of 46 years old and 73% identifying as women. In the bivariate analysis, younger age, higher education, pre-COVID-19 employment, male sex, gay/bisexual identity, and single marital status were all associated with increased intention to be vaccinated. Increasing numbers of risk factors for poorer outcomes from COVID-19 (i.e., aged 65 and older, comorbidities) were not associated with vaccine intention.

**Table 1.**
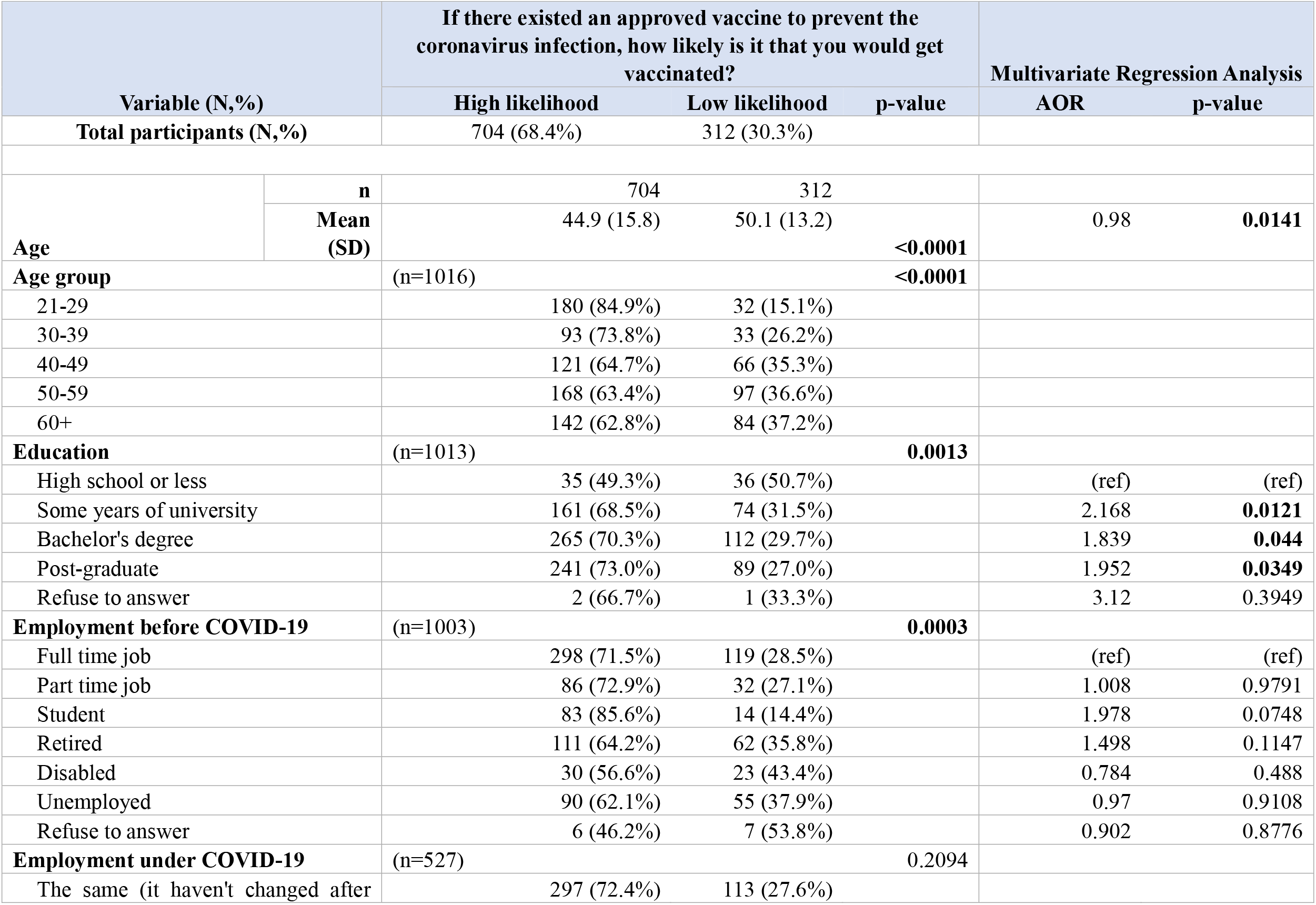
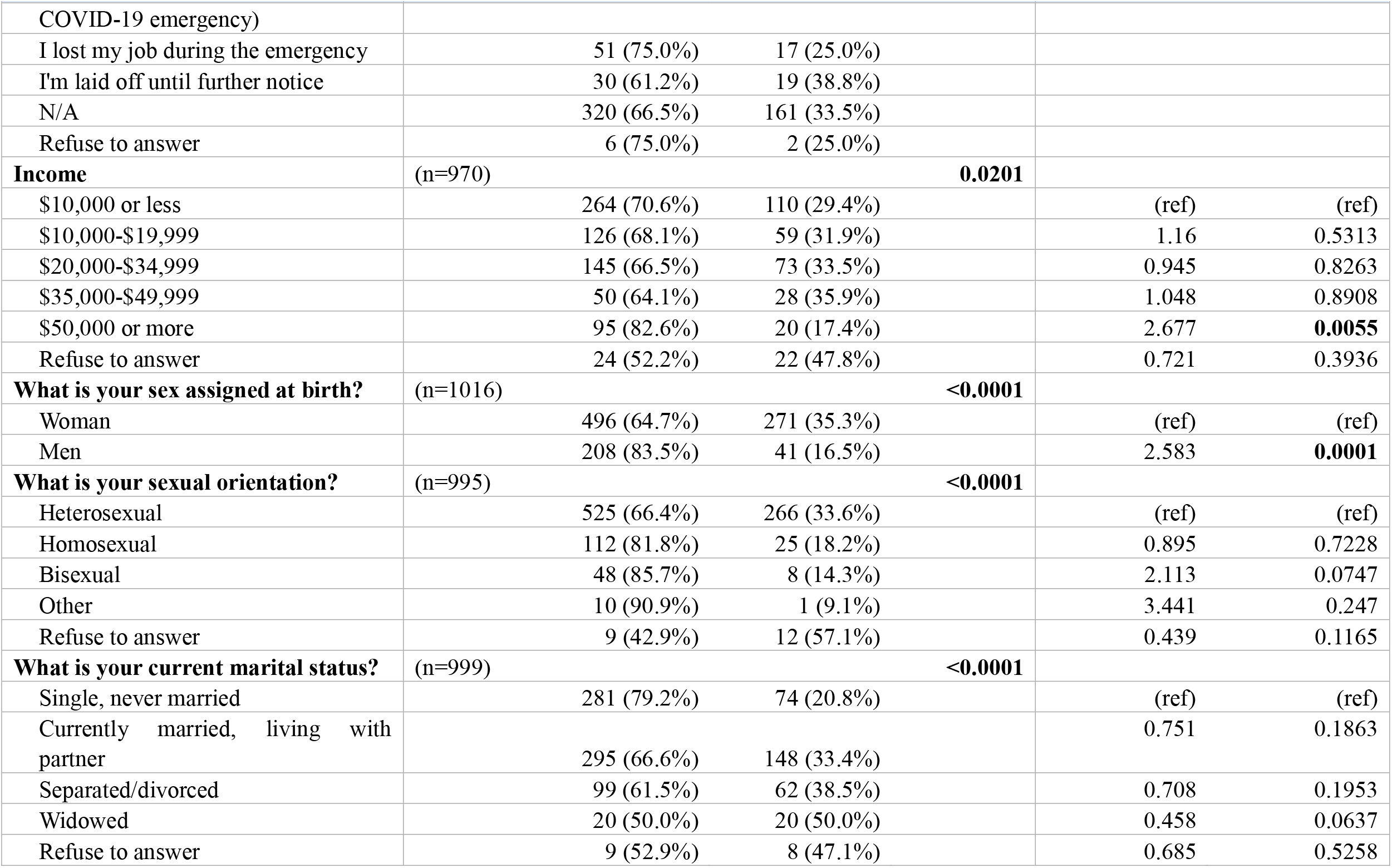
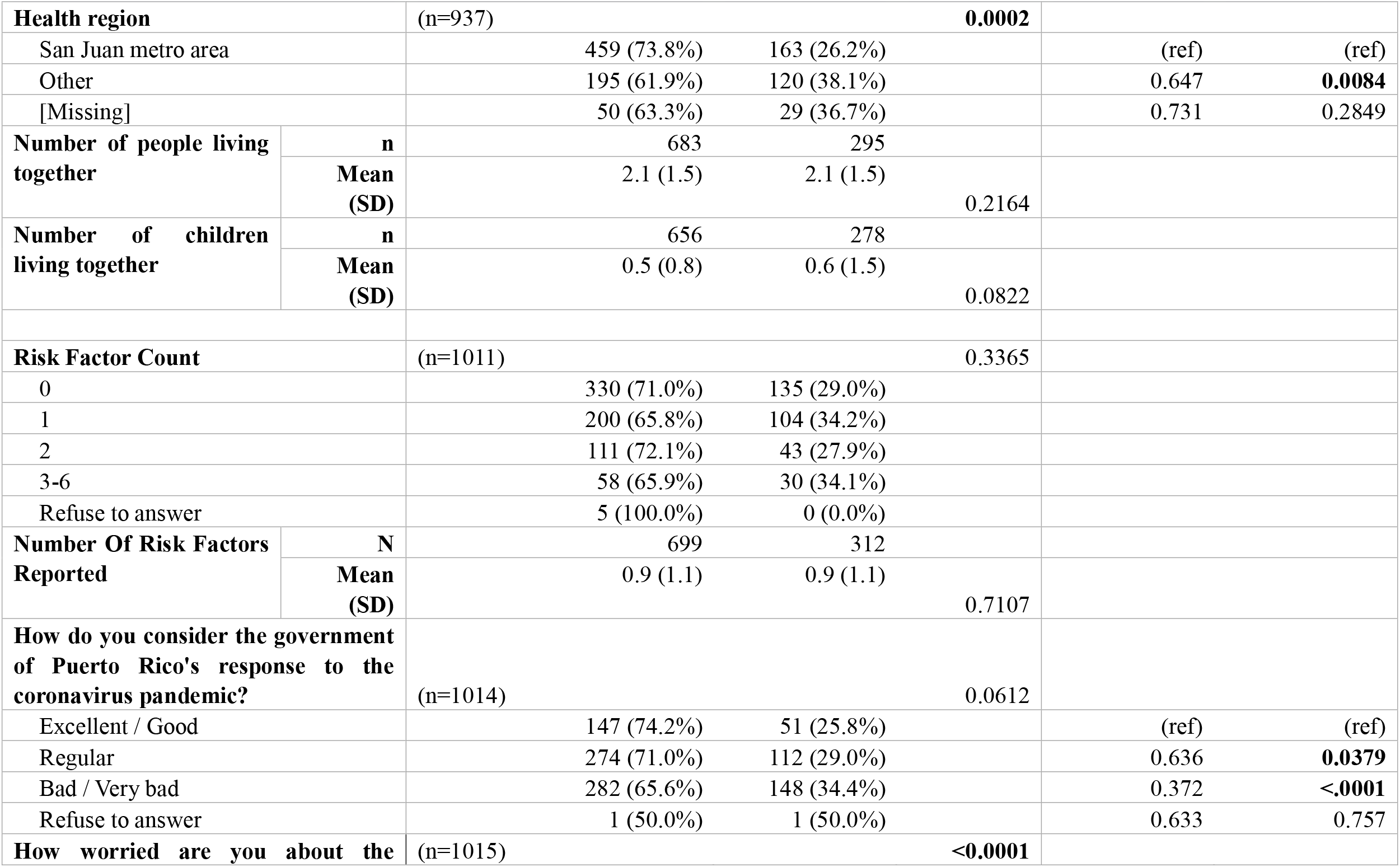
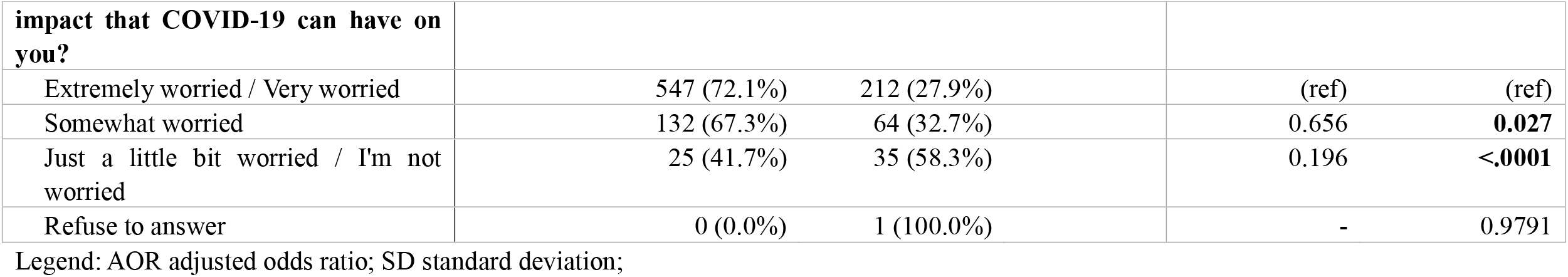
Bivariate and Multivariate Analyses of Socio-Demographic Characteristics with Vaccine Acceptability.

| Variable (N,%) |  | If there existed an approved vaccine to prevent the coronavirus infection, how likely is it that you would get vaccinated? |  |  | Multivariate Regression Analysis |  |
| --- | --- | --- | --- | --- | --- | --- |
|  |  | High likelihood | Low likelihood | p-value | AOR | p-value |
| **Total participants (N,%)** |  | 704 (68.4%) | 312 (30.3%) |  |  |  |
| **Age** | **n** | 704 | 312 |  |  |  |
|  | **Mean (SD)** | 44.9 (15.8) | 50.1 (13.2) | **<0.0001** | 0.98 | **0.0141** |
| **Age group** |  | (n=1016) |  | **<0.0001** |  |  |
| 21-29 |  | 180 (84.9%) | 32 (15.1%) |  |  |  |
| 30-39 |  | 93 (73.8%) | 33 (26.2%) |  |  |  |
| 40-49 |  | 121 (64.7%) | 66 (35.3%) |  |  |  |
| 50-59 |  | 168 (63.4%) | 97 (36.6%) |  |  |  |
| 60+ |  | 142 (62.8%) | 84 (37.2%) |  |  |  |
| **Education** |  | (n=1013) |  | **0.0013** |  |  |
| High school or less |  | 35 (49.3%) | 36 (50.7%) |  | (ref) | (ref) |
| Some years of university |  | 161 (68.5%) | 74 (31.5%) |  | 2.168 | **0.0121** |
| Bachelor's degree |  | 265 (70.3%) | 112 (29.7%) |  | 1.839 | **0.044** |
| Post-graduate |  | 241 (73.0%) | 89 (27.0%) |  | 1.952 | **0.0349** |
| Refuse to answer |  | 2 (66.7%) | 1 (33.3%) |  | 3.12 | 0.3949 |
| **Employment before COVID-19** |  | (n=1003) |  | **0.0003** |  |  |
| Full time job |  | 298 (71.5%) | 119 (28.5%) |  | (ref) | (ref) |
| Part time job |  | 86 (72.9%) | 32 (27.1%) |  | 1.008 | 0.9791 |
| Student |  | 83 (85.6%) | 14 (14.4%) |  | 1.978 | 0.0748 |
| Retired |  | 111 (64.2%) | 62 (35.8%) |  | 1.498 | 0.1147 |
| Disabled |  | 30 (56.6%) | 23 (43.4%) |  | 0.784 | 0.488 |
| Unemployed |  | 90 (62.1%) | 55 (37.9%) |  | 0.97 | 0.9108 |
| Refuse to answer |  | 6 (46.2%) | 7 (53.8%) |  | 0.902 | 0.8776 |
| **Employment under COVID-19** |  | (n=527) |  | 0.2094 |  |  |
| The same (it haven't changed after |  | 297 (72.4%) | 113 (27.6%) |  |  |  |

| Variable (N, %) | If there existed an approved vaccine to prevent the coronavirus infection, how likely is it that you would get vaccinated? |  |  | Multivariate Regression Analysis |  |
| --- | --- | --- | --- | --- | --- |
|  | High likelihood | Low likelihood | p-value | AOR | p-value |
| COVID-19 emergency) |  |  |  |  |  |
| I lost my job during the emergency | 51 (75.0%) | 17 (25.0%) |  |  |  |
| I'm laid off until further notice | 30 (61.2%) | 19 (38.8%) |  |  |  |
| N/A | 320 (66.5%) | 161 (33.5%) |  |  |  |
| Refuse to answer | 6 (75.0%) | 2 (25.0%) |  |  |  |
| **Income** | (n=970) |  | **0.0201** |  |  |
| \$10,000 or less | 264 (70.6%) | 110 (29.4%) | | (ref) | (ref) |
| \$10,000-\$19,999 | 126 (68.1%) | 59 (31.9%) | | 1.16 | 0.5313 |
| \$20,000-\$34,999 | 145 (66.5%) | 73 (33.5%) | | 0.945 | 0.8263 |
| \$35,000-\$49,999 | 50 (64.1%) | 28 (35.9%) | | 1.048 | 0.8908 |
| \$50,000 or more | 95 (82.6%) | 20 (17.4%) | | 2.677 | **0.0055** |
| Refuse to answer | 24 (52.2%) | 22 (47.8%) |  | 0.721 | 0.3936 |
| **What is your sex assigned at birth?** | (n=1016) |  | **<0.0001** |  |  |
| Woman | 496 (64.7%) | 271 (35.3%) |  | (ref) | (ref) |
| Men | 208 (83.5%) | 41 (16.5%) |  | 2.583 | **0.0001** |
| **What is your sexual orientation?** | (n=995) |  | **<0.0001** |  |  |
| Heterosexual | 525 (66.4%) | 266 (33.6%) |  | (ref) | (ref) |
| Homosexual | 112 (81.8%) | 25 (18.2%) |  | 0.895 | 0.7228 |
| Bisexual | 48 (85.7%) | 8 (14.3%) |  | 2.113 | 0.0747 |
| Other | 10 (90.9%) | 1 (9.1%) |  | 3.441 | 0.247 |
| Refuse to answer | 9 (42.9%) | 12 (57.1%) |  | 0.439 | 0.1165 |
| **What is your current marital status?** | (n=999) |  | **<0.0001** |  |  |
| Single, never married | 281 (79.2%) | 74 (20.8%) |  | (ref) | (ref) |
| Currently married, living with partner | 295 (66.6%) | 148 (33.4%) |  | 0.751 | 0.1863 |
| Separated/divorced | 99 (61.5%) | 62 (38.5%) |  | 0.708 | 0.1953 |
| Widowed | 20 (50.0%) | 20 (50.0%) |  | 0.458 | 0.0637 |
| Refuse to answer | 9 (52.9%) | 8 (47.1%) |  | 0.685 | 0.5258 |
|  |  | High likelihood | Low likelihood | p-value | AOR | p-value |
| **Health region** |  | (n=937) |  | **0.0002** |  |  |
| San Juan metro area |  | 459 (73.8%) | 163 (26.2%) |  | (ref) | (ref) |
| Other |  | 195 (61.9%) | 120 (38.1%) |  | 0.647 | **0.0084** |
| [Missing] |  | 50 (63.3%) | 29 (36.7%) |  | 0.731 | 0.2849 |
| **Number of people living together** | **n** | 683 | 295 |  |  |  |
|  | **Mean (SD)** | 2.1 (1.5) | 2.1 (1.5) | 0.2164 |  |  |
| **Number of children living together** | **n** | 656 | 278 |  |  |  |
|  | **Mean (SD)** | 0.5 (0.8) | 0.6 (1.5) | 0.0822 |  |  |
| **Risk Factor Count** |  | (n=1011) |  | 0.3365 |  |  |
| 0 |  | 330 (71.0%) | 135 (29.0%) |  |  |  |
| 1 |  | 200 (65.8%) | 104 (34.2%) |  |  |  |
| 2 |  | 111 (72.1%) | 43 (27.9%) |  |  |  |
| 3-6 |  | 58 (65.9%) | 30 (34.1%) |  |  |  |
| Refuse to answer |  | 5 (100.0%) | 0 (0.0%) |  |  |  |
| **Number Of Risk Factors Reported** | **N** | 699 | 312 |  |  |  |
|  | **Mean (SD)** | 0.9 (1.1) | 0.9 (1.1) | 0.7107 |  |  |
| **How do you consider the government of Puerto Rico's response to the coronavirus pandemic?** |  | (n=1014) |  | 0.0612 |  |  |
| Excellent / Good |  | 147 (74.2%) | 51 (25.8%) |  | (ref) | (ref) |
| Regular |  | 274 (71.0%) | 112 (29.0%) |  | 0.636 | **0.0379** |
| Bad / Very bad |  | 282 (65.6%) | 148 (34.4%) |  | 0.372 | **<.0001** |
| Refuse to answer |  | 1 (50.0%) | 1 (50.0%) |  | 0.633 | 0.757 |
| **How worried are you about the** |  | (n=1015) |  | **<0.0001** |  |  |

| Variable (N,%) | If there existed an approved vaccine to prevent the coronavirus infection, how likely is it that you would get vaccinated? |  |  | Multivariate Regression Analysis |  |
| --- | --- | --- | --- | --- | --- |
|  | High likelihood | Low likelihood | p-value | AOR | p-value |
| **impact that COVID-19 can have on you?** |  |  |  |  |  |
| Extremely worried / Very worried | 547 (72.1%) | 212 (27.9%) |  | (ref) | (ref) |
| Somewhat worried | 132 (67.3%) | 64 (32.7%) |  | 0.656 | **0.027** |
| Just a little bit worried / I'm not worried | 25 (41.7%) | 35 (58.3%) |  | 0.196 | **<.0001** |
| Refuse to answer | 0 (0.0%) | 1 (100.0%) |  | - | 0.9791 |

Legend: AOR adjusted odds ratio; SD standard deviation;

No significant relationship was observed between pre-COVID-19 employment, sexual orientation, and marital status, and intention to vaccinate after adjustment (Table 1). However, male respondents who were younger and had higher educational attainment reported higher intention to vaccinate. Respondents who had a yearly income of $50,000 or more had significantly higher intention to vaccinate (POR=2.4, p=0.0096) than those who earned $10,000 or less. Respondents living outside of the San Juan metro region had significantly lower intention to vaccinate when compared to respondents who lived in the metro region (POR=0.647, p=0.0082). Respondents who were less likely to get vaccinated expressed less worry about the impact of COVID-19 and perceived the Puerto Rican government’s response to be deficient.

Respondents who reported a high degree of confidence in one’s ability to evaluate the credibility of information about COVID-19 reported higher intention to vaccinate (Table 2). National and international health organizations (WHO, CDC) were identified as the most reliable sources for COVID-19-related information for respondents with a high likelihood of getting vaccinated (75%), followed by healthcare professionals (70%) and news programs (51%). While these sources were also selected as the most reliable among respondents with a low likelihood of vaccination, healthcare professionals and national/international health organizations were trusted at significantly lower rates than those intending to vaccinate (59% and 56%, respectively).

**Table 2.** Covid-19 Knowledge Acquisition and Perceptions with Vaccine Acceptability.

| Variable (N,%) | If there existed an approved vaccine to prevent the coronavirus infection, how likely is it that you would get vaccinated? |  |  |
| --- | --- | --- | --- |
|  | High likelihood | Low likelihood | p-value |
| **How confident are you in your ability to determine if the information you are receiving about Covid-19 is correct?** | (n=1007) |  | **0.0011** |
| Super confident | 288 (76.2%) | 90 (23.8%) |  |
| Somewhat confident | 366 (65.5%) | 193 (34.5%) |  |
| Not confident at all | 44 (62.9%) | 26 (37.1%) |  |
| Refuse to answer | 6 (66.7%) | 3 (33.3%) |  |
| **What sources do you trust to obtain reliable information regarding COVID-19?\*** |  |  |  |
| (select all, n=1016) |  |  |  |
| Social media | 303 (43.0%) | 121 (38.8%) | 0.2043 |
| TV commercials | 155 (22.0%) | 63 (20.2%) | 0.5134 |
| News programs | 358 (50.9%) | 156 (50.0%) | 0.8021 |
| Peer network | 115 (16.3%) | 34 (10.9%) | **0.0238** |
| Healthcare professionals | 495 (70.3%) | 186 (59.6%) | **0.0008** |
| National/international health organizations | 525 (74.6%) | 174 (55.8%) | **<0.0001** |
| Local health department | 349 (49.6%) | 137 (43.9%) | 0.0955 |
| Other | 39 (5.5%) | 12 (3.8%) | 0.2541 |
| I don't believe any source of information about COVID 19 | 3 (0.4%) | 11 (3.5%) | **0.0001** |
| Refuse to answer | 5 (0.7%) | 5 (1.6%) | - |

\*Note: Respondents were able to select multiple options. Percentages noted for individual information sources are calculated by dividing the number of respondents who selected each information source divided by the total number of participants in the column (either high likelihood vs. low likelihood).

## Discussion

These results, which are part of a larger survey created to evaluate the impact of COVID-19 on people living in Puerto Rico, provide insight into vaccine acceptability and intention to vaccinate amongst the participants and may be useful to current vaccination efforts in Puerto Rico and the U.S. Although the total number of risk factors for poor COVID-19 outcomes was not associated with intention to vaccinate, decreased intention observed among older participants is troubling. Several limitations should be considered when interpreting the findings of this study. As mentioned previously, vaccine acceptability tends to decrease over time following the onset of a pandemic, and responses from July of 2020 may overestimate intention to vaccinate. Also, the item assessing intention to vaccinate was posed in a hypothetical manner as no vaccine was yet available. As such, perceptions and intentions may have changed since that time, in particular with regards to media coverage of different vaccine manufacturers and vaccines currently approved or in development.

## Conclusions

These findings highlight the importance of considering sociodemographic characteristics identified with low intention to vaccinate when designing public messaging related to increasing COVID-19 vaccinations. In particular, older individuals earning less than $50,000 a year and those living in rural areas may be at particular risk of not being vaccinated. Furthermore, in light of reported perceptions of the government’s response to the COVID-19 crisis, communication initiatives should consider efforts to promote vaccinations via trusted sources of information, particularly among those who are less likely to get vaccinated. Communication campaigns may benefit from using established guidance from the CDC and WHO, and by being delivered by local healthcare professionals and through news outlets to ensure success.

## Data Availability

Due to confidentiality agreements, supporting data can only be made available to bona fide researchers subject to a non-disclosure agreement. For details of the data and how to request access contact Dr. Carlos E. Rodriguez-Diaz

## Notes

**Financial Disclosure:** The work of Juan Pablo Zapata was supported by the National Center for Advancing Translational Sciences, National Institutes of Health, through grant numbers UL1TR001436 and 1TL1TR001437. No other financial disclosures were reported by the authors of this paper.

### Competing Interest Statement

The authors have declared no competing interest.

### Funding Statement

The work of Juan Pablo Zapata was supported by the National Center for Advancing Translational Sciences, National Institutes of Health, through grant numbers UL1TR001436 and 1TL1TR001437. No other financial disclosures were reported by the authors of this paper.

### Author Declarations

Ethical approval for the study protocol was provided by the Human Research Subjects Protection Office at the University of Puerto Rico-Medical Sciences Campus (Protocol #A9650220).

